# Risk-benefit analysis of the AstraZeneca COVID-19 vaccine in Australia using a Bayesian network modelling framework

**DOI:** 10.1101/2021.09.30.21264337

**Authors:** Colleen L Lau, Helen J Mayfield, Jane E Sinclair, Samuel J Brown, Michael Waller, Anoop K Enjeti, Andrew Baird, Kirsty Short, Kerrie Mengersen, John Litt

## Abstract

Thrombosis and Thromobocytopenia Syndrome (TTS) has been associated with the AstraZencea (AZ) COVID-19 vaccine. Australia has reported low TTS incidence of <3/100,000 after the first dose, with case fatality rate (CFR) of 5-6%. Risk-benefit analysis of vaccination has been challenging because of rapidly evolving data, changing levels of transmission, and age-specific variation in rates of TTS, COVID-19, and CFR. We aim to optimise risk-benefit analysis by developing a model that enables inputs to be updated rapidly as evidence evolves. A Bayesian network was used to integrate local and international data, government reports, published literature and expert opinion. The model estimates probabilities of outcomes under different scenarios of age, sex, low/medium/high transmission (0.05%/0.45%/5.76% of population infected over 6 months), SARS-CoV-2 variant, vaccine doses, and vaccine effectiveness. We used the model to compare estimated deaths from vaccine-associated TTS with i) COVID-19 deaths prevented under different scenarios, and ii) deaths from COVID-19 related atypical severe blood clots (cerebral venous sinus thrombosis & portal vein thrombosis). For a million people aged ≥70 years where 70% received first dose and 35% received two doses, our model estimated <1 death from TTS, 25 deaths prevented under low transmission, and >3000 deaths prevented under high transmission. Risks versus benefits varied significantly between age groups and transmission levels. Under high transmission, deaths prevented by AZ vaccine far exceed deaths from TTS (by 8 to >4500 times depending on age). Probability of dying from COVID-related atypical severe blood clots was 58-126 times higher (depending on age and sex) than dying from TTS. To our knowledge, this is the first example of the use of Bayesian networks for risk-benefit analysis for a COVID-19 vaccine. The model can be rapidly updated to incorporate new data, adapted for other countries, extended to other outcomes (e.g., severe disease), or used for other vaccines.

**HIGHLIGHTS:**

- AZ vaccination risk-benefit analysis must consider age/community transmission level
- AZ vaccine benefits far outweigh risks in older age groups and during high transmission
- AZ vaccine-associated TTS lower fatality than COVID-related atypical blood clots
- Bayesian networks utility for risk-benefit analysis of rapidly evolving situations
- BNs allow integrating multiple data sources when large datasets are not available

## 1. INTRODUCTION

The AstraZeneca ChAdOx1 (AZD1222) COVID-19 vaccine (AZ vaccine) has been widely used globally, with one billion doses released in over 170 countries by August 2021 [1]. The vaccine is highly effective against symptomatic infection, serious illness and death from COVID-19 [2-4]. In March 2021, reports of AZ vaccine-associated Thrombosis with Thrombocytopenia Syndrome (TTS) began emerging from multiple countries, including Norway [5], Canada [6], the U.K. [7], and Australia [8], and appeared to be more common in younger age groups [8]. Although the incidence of TTS was very low (<3/100,000 first doses in any age group in Australia), initial reports of high case fatality rates (CFR) (44% in Germany [9] and 18% in United Kingdom [7]) prompted some countries to recommend alternative COVID-19 vaccines in younger persons.

In Australia, a change in recommendation was made in April 2021 to preference the Pfizer/BioNTech (BNT162b2) COVID-19 vaccine over the AZ vaccine in those aged <50 years [10]. In June 2021, after the death of a 52-year-old woman, the recommendation was revised to preference the Pfizer vaccine over the AZ vaccine in those aged <60 years [11, 12]. The AZ vaccine-associated TTS cases and deaths received significant media attention in Australia, resulting in increased vaccine reluctance, and uncertainty amongst the public and clinicians about risks versus benefits of vaccination [13]. The only other COVID-19 vaccine available in Australia at the time (and up to early September 2021) was the Pfizer vaccine, but limited supplies meant that those choosing the Pfizer vaccine had to wait weeks or months to be vaccinated.

Initial risk assessments and decisions about age-based recommendations for the AZ vaccine in Australia were made at a time when there was almost no local community transmission of COVID-19. Unfortunately, since late June 2021, outbreaks have occurred in multiple states and territories, resulting in prolonged lockdowns of millions of people in New South Wales (NSW), Victoria (VIC), and the Australian Capital Territory (ACT) [14]. Therefore, the risks versus benefits of the AZ vaccine changed dramatically over a few weeks, and those aged <60 years have been advised to see a doctor to help them make an individual risk assessment.

On 29 June 2021, the Australian Government produced a document on ‘Weighing up the potential benefits against the risk of harm from COVID-19 Vaccine AstraZeneca’ for clinicians to help patients make informed decisions [15]. The document provides helpful information for each age group such as the risk of TTS versus the number of deaths, intensive care unit (ICU) admissions, and hospitalisations prevented by vaccination under low, medium, and high levels of transmission. However, calculations of risks versus benefits changed rapidly because of many factors, including evolving incidence and CFR of TTS (both in Australia and internationally), changing levels of community transmission, changing age distribution of cases associated with the arrival of the delta variant (the predominant variant in Australia since June 2021), and new data on vaccine effectiveness against the delta variant. These and other factors that influence risk-benefit analysis are likely to continue to evolve at a rapid pace.

To address the challenge of providing up-to-date risk-benefit analysis of COVID-19 vaccination, one of the authors (JL) suggested to the Immunisation Coalition (https://www.immunisationcoalition.org.au/) that they support the development of the COVID-19 Risk Calculator (CoRiCal), a set of tools designed to facilitate shared decision making between clinicians and patients. The Immunisation Coalition is a not-for-profit organization with representation from clinical and other professional groups interested in promoting vaccination best practice in Australia. CoRiCal aims to support informed risk assessment by providing relevant and accurate estimates of the risks and benefits of COVID-19 vaccines, taking into account locally relevant factors.

This paper describes the first CoRiCal model, focused on risk-benefit analysis of the AZ vaccine. The model was developed using a Bayesian network (BN), which allows the model’s inputs to be updated easily as the outbreak situation changes and as scientific evidence evolves. The model aims to provide more precise risk-benefit analysis of AZ vaccination by comparing the risk of death from vaccine-associated TTS versus the risk death from i) COVID-19 infection, and ii) atypical severe blood clots (cerebral venous sinus thrombosis [CVST] and portal vein thrombosis [PVT]) related to COVID-19 infection. The model enables scenario analysis based on age, sex, current transmission intensity, predominant SARS-CoV-2 variant, number of AZ vaccine doses received, and vaccine effectiveness against symptomatic infection and death. The model could also be extended for outcomes other than death (e.g., ICU admission, long COVID), other vaccines, other adverse events following immunization (AEFI), or adapted for use in other countries.

## 2. METHODS

### 2.1. Bayesian networks

BNs are acyclic graphical models that explicitly represent relationships between variables in terms of conditional probabilities [16]. Variables are represented by nodes, each categorized into several possible states (e.g., yes/no, high/low, age groups). Links (or edges) represent the relationships between nodes and point in the direction from a parent node (independent variable) to a child node (dependent variable) (Figure 1). Nodes are quantified using probability tables that define the probability of a node being in a given state, either based on some prior distribution (for nodes with no parent nodes) or conditional on the states of all parent nodes i.e., conditional probability tables (CPTs).

**Figure 1.**
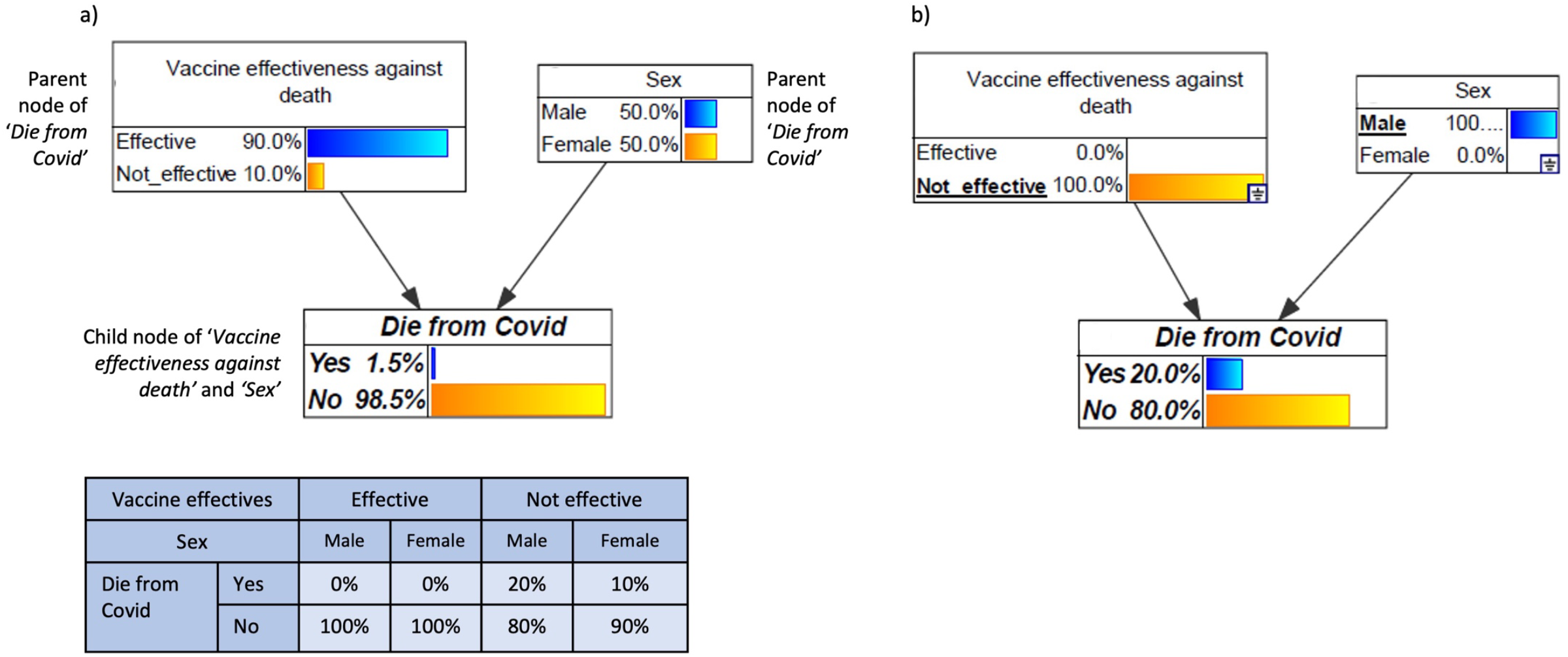
Example Bayesian network showing three nodes (white boxes), links (arrows), and conditional probability table (CPT) for *Die from Covid* node (blue table). Data in figure are fictitious and for illustrative purposes only. The model assumes 90% vaccine effectiveness against death and even distribution of males or females in the population; these numbers are entered as priors because the nodes do not have parents. The model assumes case fatality rate of 20% in males and 10% in females. The CPT for *Die from Covid* shows that if the vaccine is effective, probability of dying is 0% for both genders. If the vaccine is not effective, the probability of dying is 20% for males and 10% and females. Fig 1a) shows the BN in its default state; if the vaccine was 90% effective, we expect an overall 1.5% chance of dying from COVID (e.g., in a population of 1000 people, the vaccine was not effective in 100 people. Of these 100 people, we estimate 10 deaths out of 50 males, and 5 deaths out of 50 females, i.e., total of 15 deaths out of 1000 people, or 1.5%). Fig 1b) shows an example scenario analysis of ‘what is the chance of a male dying if the vaccine was not effective?’ Selected states in each node are underlined, and the model updates the chance of dying from COVID to 20% under this scenario.

BN modelling frameworks offer many advantages relevant to the problem at hand. Firstly, BNs provide visual and transparent representation of relationships between variables and the probabilistic assumptions between each variable. The graphical structure enables users to understand how the system has been modelled, clearly shows the underlying assumptions, how model outputs were derived, and enables interactive scenario analysis. Secondly, CPTs can be derived from a range of data formats, allowing relevant data to be integrated from diverse sources including studies, reports, and expert opinion. Thirdly, the probabilistic framework of BNs is well suited to risk analysis where the uncertainty surrounding estimates can be tested through sensitivity analysis for a range of plausible inputs. Finally, model assumptions and CPTs can be updated easily as more information become available or as the situation evolves. Rapid adaptability is particularly important for modelling rapidly evolving situations such as outbreaks.

BNs have been used for a variety of COVID-19 models, including modelling transmission and outbreak response [17, 18], decision making [19], risk analysis [20, 21], risk assessment and contact tracing [22, 23], interpretation of SARs-CoV-2 test results [24], and predictive diagnosis [25]. However, to our knowledge, BNs have not yet been applied for risk-benefit analysis of COVID-19 vaccines.

### 2.2. Model design

Our BN model structure was based on the best available scientific evidence from multiple sources. Evidence was integrated using facilitated expert elicitation, with subject matter experts (JL, AB, KRS, AKE, CLL, Thrombosis & Haemostasis Society of Australia and New Zealand [THANZ]) working closely with experienced BN modellers (HJM, KM, CLL). The four subject matter experts included clinicians and researchers with expertise in infectious disease epidemiology, virology, thrombosis, general practice, and public health. The experts defined the scope of the model, key outcome variables of interest, and major predictor variables (inputs) that influenced the outcomes. Experts reviewed published literature and reports to determine whether there was sufficient evidence to include each predictor variable in the model. Predictors were linked to outcomes if there were sufficient evidence and data to quantify the conditional probabilities between them.

Based on the discussions between the experts and modellers, and evidence collected by the experts, a draft BN model structure was derived by the modellers, then reviewed and refined with the team over several iterations of facilitated discussions. To parameterize the model, questionnaires were developed to help experts structure the systematically collected data into CPTs. Variables were not linked if there were only weak quantitative relationships between them because the links would have little impact on model predictions. Data were updated as new evidence became available during the model development process from July-September 2021. External consistency was considered during model design so that predictions were consistent with other commonly used information sources such as the Australian Technical Advisory Group on Immunisation (ATAGI) and the Therapeutic Goods Administration (TGA), e.g., using the same age groups and definitions of low/medium/high community transmission intensity (equivalent to 0.05%/0.45%/5.76% of population infected over 6 months) [15].

### 2.3. Atypical severe blood clots

TTS is a rare and atypical form of blood clotting associated paradoxical thrombosis and low platelets. TTS is immunologically mediated, and can be trigged by medications (e.g., heparin) or vaccines (e.g., AZ vaccine) [26]. TTS is diagnosed on the basis of clinical criteria and includes atypical severe blood clots such as CVST and splanchnic vein thrombosis (SVT) [27, 28], which have also been reported in COVID-19 patients [29, 30]. CVST and SVT are distinct from the common types of venous thromboembolism such as deep vein thrombosis or pulmonary embolus, or other types of thrombocytopenia and have the highest risk of mortality in this group. SVT includes portal vein thrombosis (PVT), Budd Chiari Syndrome and mesenteric vein thrombosis. The data included in the model focused on PVT as it is the most common type of SVT, and has been specifically reported post-COVID, enabling a direct comparison. Our model therefore focused on comparing the risk of AZ vaccine-associated TTS with the risk of CVST and PVT in COVID-19 patients.

### 2.4. Data sources

While the assumptions and model structure are defined by experts, the CPTs were based on empirical data. Experts compiled evidence from peer-reviewed literature, government websites and reports, and through discussion with external clinical experts (e.g., haematologists regarding the evidence for AZ vaccine-associated TTS, and background rates of CVST and PVT). Official data from Australian authorities were used wherever possible (e.g., local data on AZ vaccine-associated TTS). Otherwise, data were obtained from other robust and publicly available sources (e.g., background rates of CVST and PVT). Data from different sources were harmonised for the CPTs, e.g., summarising data to match the age groups used in the BN. Data analyses were conducted for some variables to convert them into probabilities for the CPTs, e.g., converting incidence of COVID-19 into probability of infection over 6 months for the *Intensity of local transmission* node. Table 1 [31-40] and Appendix A provide a summary of data sources, model assumptions, and rationale.

**Table 1.** Summary of data sources, assumptions, and prior distributions for a Bayesian network to assess risks versus benefits of the AstraZeneca COVID-19 vaccine.

| Model inputs | Data sources, assumptions, rationale | Reference |
| --- | --- | --- |
| Age distribution of infections from delta variant | NSW COVID-19 case data from 1/6/2021 to 13/8/2021 were used to provide estimates of age distribution of infections from delta variant. Case data published daily by NSW Health, for the following age categories: 0-19, 5-year age groups from 20-69 years, and 70+. For cases in the 0-19 age group, assumed that 40% were aged 0-9, and 60% aged 10-19 (based on age distribution of cases reported by NNDSS). Date range used was selected to reflect the first 6 weeks of delta outbreak, when vaccination coverage was relatively low. No significant change in age distribution of cases to 29/8/2021. See Appendix A, Table A1. | 31,32 |
| Age distribution of infections from alpha/wild variants | COVID-19 cases reported in Australia from January to December 2020 were used to provide estimates of age distribution from alpha/wild variants. Data sourced from National Notifiable Diseases Surveillance System. See Appendix A, Table A2. | 14,33 |
| Case fatality rates of COVID-19 cases | COVID-19 cases reported in Australia from January 2020 to 13/8/2021 were used to provide estimates of age-specific case fatality. Data sourced from National Notifiable Diseases Surveillance System. See Appendix A, Table A3. | 14,33 |
| Community transmission levels | Chance of infection over 6 months calculated for different levels of community transmission. See Appendix A, Table A4.<br>Definitions of low, medium, and high transmission as defined by ATAGI document 'Weighing up the potential benefits and risk of harm from COVID-19 Vaccine AstraZeneca'. Low – similar to first wave in Australia (equivalent to 0.05% of population infected over 6 months). Medium – similar to second wave in VIC in 2020 (equivalent to 0.045% of population infected over 6 months). High – similar to Europe in January 2021 (equivalent to 5.76% of population infected over 6 months).<br>Also included transmission scenarios equivalent to: zero transmission; 1% and 2% chance of infection over 6 months; 200 cases/day and 1000 cases/day in NSW; 1000 cases/day in VIC; 1000 cases/day in QLD. Other transmission levels can be added to model. | 15 |
| Chance of infection by age group | Chance of infection differed between age groups and by variants. Calculated chance of infection by age group if overall community transmission of 1%. Calculations based on age distribution of infections from delta and alpha/wild variants, and age distribution of Australian population. See Appendix A, Table A5. |  |
| Vaccine effectiveness against symptomatic infection | Delta variant (ATAGI recommended data used in Doherty transmission model):<br>• 33% effective after 1<sup>st</sup> dose<br>• 61% effective after 2<sup>nd</sup> dose<br>(Note: Public Health England estimates effectiveness of 45% after 1<sup>st</sup> dose, and 70% after 2<sup>nd</sup> dose) | 34,35 |
|  | Alpha variant (from Vaccine effectiveness Expert Panel, Public Health England):<br>• 60% effective after 1<sup>st</sup> dose<br>• 80% effective after 2<sup>nd</sup> dose |  |
| Vaccine effectiveness against death | Delta variant (ATAGI recommended data used in Doherty transmission model):<br>• 69% effective after 1<sup>st</sup> dose<br>• 90% effective after 2<sup>nd</sup> dose<br>(Note: Public Health England estimates effectiveness of 80% after 1<sup>st</sup> dose, and 95% after 2<sup>nd</sup> dose).<br>Alpha variant (from Vaccine effectiveness Expert Panel, Public Health England):<br>• 80% effective after 1<sup>st</sup> dose<br>• 95% effective after 2<sup>nd</sup> dose | 34,35 |
| Thrombosis and Thrombocytopenia Syndrome (TTS) after AZ vaccine | Model uses data reported by ATAGI update following weekly COVID-19 meeting on 25/8/2021.<br>Estimated rate per 100,000 1<sup>st</sup> dose of AZ vaccinations:<br>• Age <50: 2.5<br>• Age 50-59: 2.7<br>• Age 60-69: 1.6<br>• Age 70-79: 2.1<br>• Age ≥80: 1.6<br>• For age ≥70 in model, used rate of 1.85 (average of rates for 70-79 and ≥80).<br>Estimated rate per 100,000 after 2<sup>nd</sup> dose of AZ vaccinations: 0.18 per 100,000 (no age specific rates available).<br>Case fatality rate in Australia ~5% (noting that higher rates reported in UK ~18%).<br>For sensitivity analysis, data from ATAGI reports on 1/9/2021, 8/9/2021, and 15/9/2021 were used. | 36–39 |
| Background rates of atypical venous thrombotic disorders | Background rates (in population not infected with and not vaccinated for COVID-19) of atypical venous thrombotic disorder (CVST and PVT) over 6 weeks were calculated for each age group to provide a comparison with chance of TTS after AZ vaccine.<br>CVST data from Kristoffersen *et al.*<br>• Age-specific rates per million population per year:<br>○ Age <20: 10.8<br>○ Age 20-49: 18.0<br>○ Age 50-69: 21.1<br>○ Age ≥70: 20.7<br>• Case fatality of 7% for all age groups.<br>• Assumed equal rates in males and females. | 27,28 |

|  | PVT data from Ageno *et al.*<br>• Age-specific rates per million population per year:<br>○ Age <20: 0<br>○ Age 20-29: 5.5<br>○ Age 30-39: 7.25<br>○ Age 40-49: 15.75<br>○ Age 50-59: 25.5<br>○ Age 60-69: 49.5<br>○ Age ≥70: 55.125<br>• Case fatality of 27.2% for all age groups.<br>• Assumed equal rates in males and females. |  |
| --- | --- | --- |
| Atypical venous thrombotic disorders associated with COVID-19 infection | Rates of CVST and PVT in COVID-19 cases from a retrospective cohort study using data primarily from the USA.<br>CVT:<br>• Cases per million COVID-19 infections:<br>○ Male: 28.87<br>○ Female: 54.20<br>• Case fatality 17.4% for both sexes.<br>• Assumed same rates for all age groups.<br>PVT:<br>• Cases per million COVID-19 infections:<br>○ Male: 483<br>○ Female: 318<br>• Case fatality 19.9% for both sexes.<br>• Assumed same rates for all age groups. | 30 |
| Prior distributions* | Assumptions | Reference |
| Age distribution of Australian population | Australian Bureau of Statistics. National population estimates, December 2020. See Appendix A, Table A6. | 40 |
| Gender distribution of Australian population | 50% male, 50% female |  |
| Variants | 90% delta, 10% alpha/wild |  |
| Vaccine coverage | 70% received 1<sup>st</sup> dose, 35% received 2<sup>nd</sup> dose |  |

ATAGI=Australian Technical Advisory Group on Immunisation; CVST=Cerebral venous sinus thrombosis; PVT=Portal vein thrombosis. \*Note that prior distributions do not affect results of scenario analysis but enables the model to provide population-level estimates. Assumptions can be changed as the situation evolves.

The model includes default prior distributions for age group (using age distribution of Australia), gender (50% male, 50% female), SARS CoV-2 variants (90% delta, 10% alpha/ancestral virus), and vaccine coverage (70% of population received first dose, 35% received two doses). The prior distributions do not affect the results of scenario analyses, e.g., if delta was selected in the variants node, the model outputs relate to delta only regardless of the prior distribution of variants entered into the model. Prior distributions can be changed to model other scenarios, e.g., different distributions of variants, and different levels of vaccine coverage.

### 2.5. Model validation

The final model was assessed by subject matter experts and the modelling team together, to determine if the structure, variables, and assumptions reflected current knowledge and evidence. The model’s predictive ability was not validated against a dataset because model parameters were not learned from data. Rather, we validated each section of the model by defining a range of scenarios and manually assessed whether model outputs were consistent with our assumptions. Manual quantitative validations were conducted by a subject matter expert/BN modeller (CLL), a mathematician/statistician/BN modeller (KM) and a biostatistician (MW). Scenarios used for manual validations are provided in Appendix B. All authors evaluated the biological plausibility of estimates, e.g., for COVID-19 patients, the probability of dying from COVID-19 related atypical blood clots should be lower than the total probability of dying from COVID-19.

### 2.6. Sensitivity analysis

To determine how often model assumptions need to be updated, sensitivity analysis was conducted for three predictor variables that were considered most likely to change over time. We examined actual changes in the reported incidence of AZ vaccine-associated TTS cases and deaths in Australia in August-September 2021. We also examined i) plausible differences in age-specific CFR for COVID-19 in Australia (e.g., if higher CFR with new variants or because of an overwhelmed health system, and ii) plausible reductions in vaccine effectiveness against symptomatic COVID-19 infection and death.

#### Incidence and CFR of vaccine-associated TTS

From July-September 2021, reported incidence of AZ vaccine-associated TTS and related deaths in Australia fluctuated by week because of low numbers. We examined ATAGI reports from 25/8/2021, 1/9/2021, 8/9/2021, and 15/9/2021 [36-39] to determine how fluctuations in data affected our model predictions of age-specific TTS-related deaths over these weeks.

#### Age-specific CFR for COVID-19 in Australia

By 31/8/2021, COVID-19 CFR in Australia were very low for younger age groups, with less than five deaths during the entire pandemic in each male/female subgroups in those aged 0-9, 10-19, 20-29, 30-39, and 40-49 years. We examined changes in CFR if (theoretically) there was one extra death or five extra deaths in each age-sex subgroup, and the potential impact of these changes on model predictions on COVID-19 related deaths.

#### Vaccine effectiveness against symptomatic infection and death

We examined the sensitivity of model outputs to reduced vaccine effectiveness against the delta variant using theoretical assumptions of 5% and 10% reduction in effectiveness against both symptomatic infection and death.

## 3. RESULTS

### 3.1. Model description

The final BN structure (Figure 2) synthesises the assumptions regarding probabilistic relationships between variables and outcomes. Table 2 provides a summary of all nodes (n1 to n20) and their parent/child relationships. The model was designed to predict five outcomes:

**Table 2.**
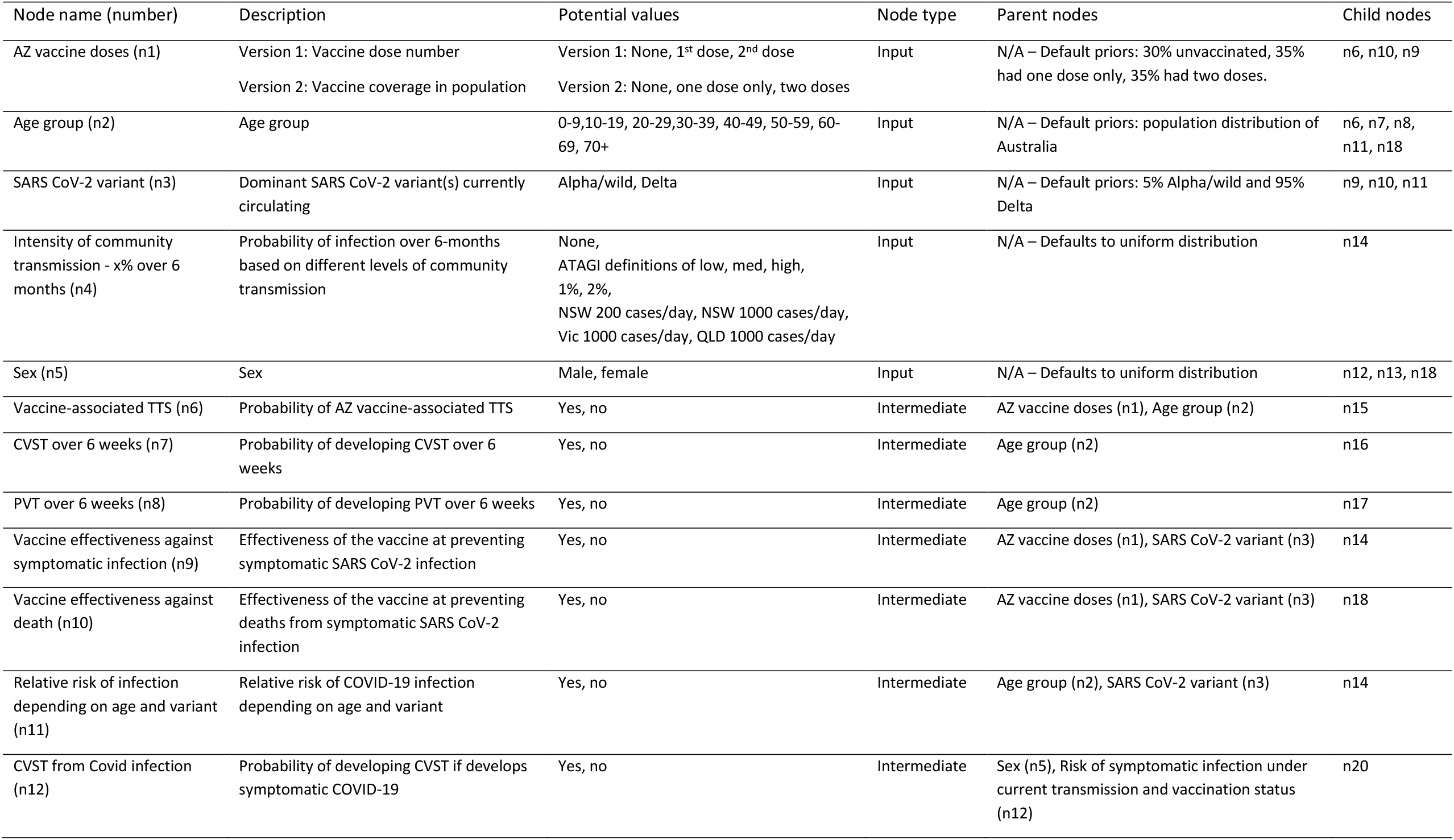
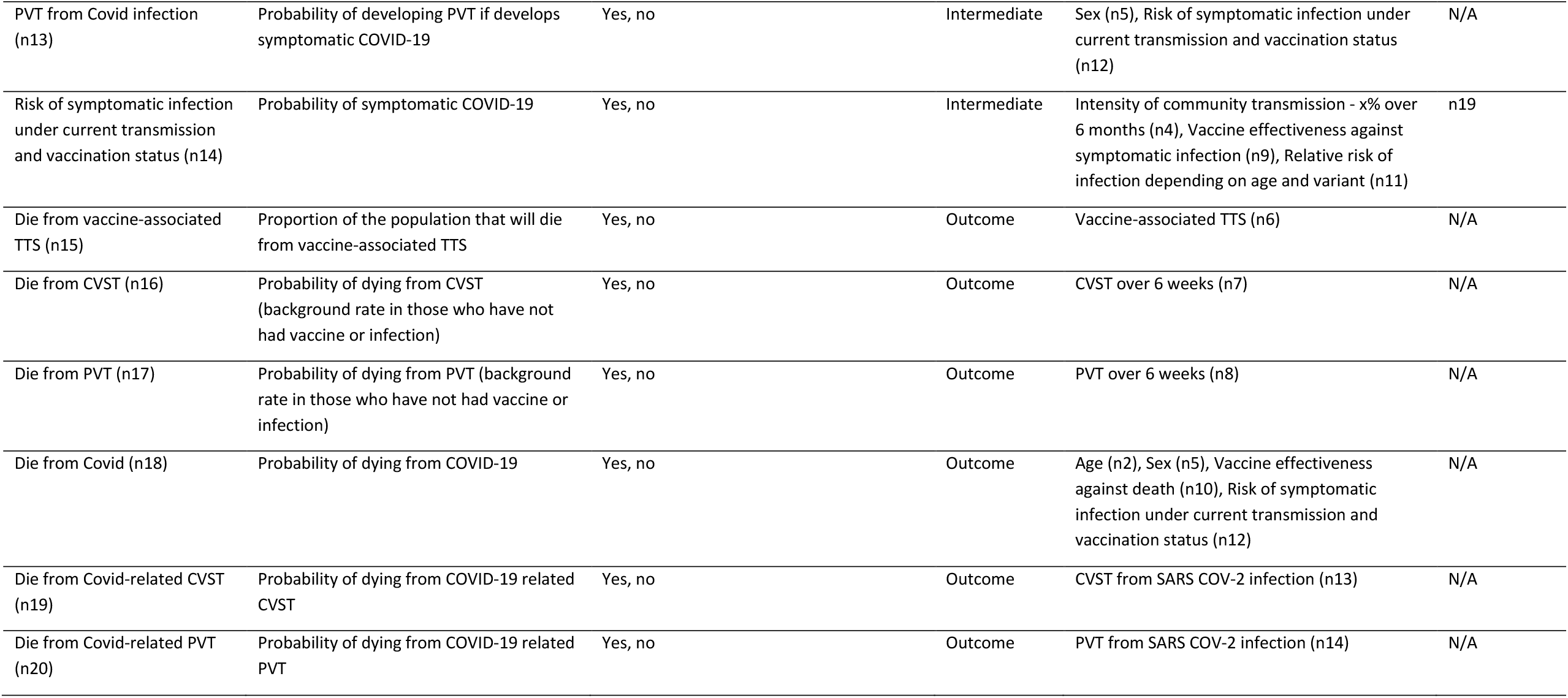
Summary of nodes and relationships between nodes in a Bayesian network for assessing risks versus benefits of the AstraZeneca COVID-19 vaccine.

**Figure 2.**
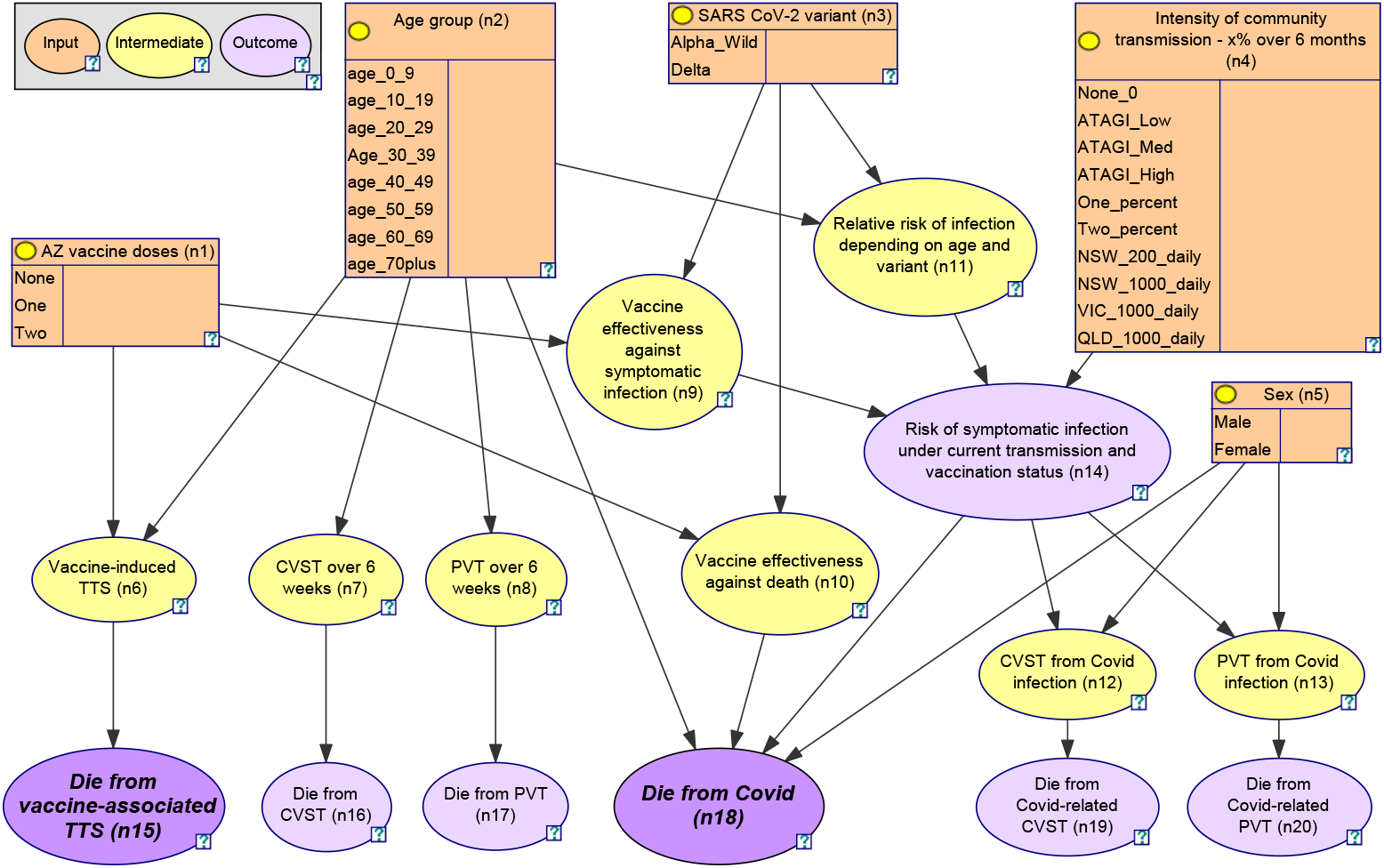
Bayesian network for assessing risks versus benefits of the AstraZeneca COVID-19 vaccine.

i. Probability of dying from AZ vaccine-associated TTS (n6) – depending on age (n2), 1^st^ or 2^nd^ dose of AZ vaccine (n1);
ii. Background probability of deaths from CVST and PVT (in those who have not had AZ vaccine or COVID-19 infection) (n7, n8). Estimates were converted to probability of events over 6 weeks to enable comparison with the probability of vaccine-associated TTS, which generally occurs within 6 weeks of vaccination;
iii. Probability of symptomatic COVID-19 infection – depending on age (n2), sex (n5), variant (n3), intensity of community transmission (n4), vaccine effectiveness against symptomatic infection (n9);
iv. Probability of dying from COVID-19 (n18) – depending on age (n2), sex (n5), variant (n3), intensity of community transmission (n4), vaccine effectiveness against symptomatic infection (n9), vaccine effectiveness against death (n10); and
v. Probability of CVST and PVT deaths (n19, n20) related to COVID-19 infection – depending on sex (n5).

The BN includes five input nodes (orange) that could be used for scenario analysis based on AZ vaccine doses (n1), age (n2), sex (n5), variant (n3) and intensity of community transmission (n4). Community transmission scenarios were converted to probability of infection over six months to more readily compare the risks versus benefits of vaccination, which is expected to be effective for at least six months. Scenarios were selected based on ATAGI definitions of low/medium/high risk [15], other scenarios that users could relate to (e.g., 1000 cases/day in NSW), and general scenarios that could be adapted to any setting (e.g., 1% chance of infection over 6 months). The model includes nine intermediate nodes (yellow) where modellers can readily update evidence on incidence of TTS (n6), background incidence of CVST and PVT (n7, n8), relative risk of infection based on age and variant (n11), vaccine effectiveness (n9, n10), and incidence and CFR of COVID-19 related CVST and PVT (n12, n13).

Two versions of the model were built using different definitions of the ‘AZ vaccine doses (n1)’ node:

- Version 1: AZ vaccine doses defined as no doses, 1^st^ dose, and 2^nd^ dose. This version was used to estimate the probability of vaccine-associated TTS with each dose of vaccine.
- Version 2: AZ vaccine doses defined as no doses, received only one dose, and received both doses. This version was used to estimate the probability of deaths in the population based on vaccine coverage rates.

### 3.2. Model validation

A model walk-through indicated that subject matter experts agreed the model structure matched their understanding of the problem space in terms of the relevant predictors and relationships drawn from the external data sources. Independent manual calculations by three authors (CLL, MW, KM) of probabilities of selected outcomes were consistent with model predictions (Appendix B).

A minor anomaly was detected in the model through assessment of biological plausibility. For COVID-19 cases in younger age groups (females <30 years and males <10 years), the estimated probability of dying from COVID-19 was lower than the probability of dying from COVID-19 related atypical severe blood clots. The reason for this anomaly is that data for COVID-19 CFR in Australia were used for the CPTs, and there have been no deaths to date in these age-sex subgroups (Table A3), while data on COVID-19 related atypical blood clots were extracted from a study where data were predominantly sourced from the USA and Europe, where high CFRs could have resulted from an overwhelmed health system during the outbreak. Although the discrepancies in probabilities were extremely low (probability of dying from COVID-19 related atypical blood clots <0.0002% higher than dying from COVID-19 itself), the anomaly highlights that the model predictions should be used as broad estimates rather than exact risks.

### 3.3. Comparison of estimated deaths prevented from COVID-19 under different scenarios of community transmission intensity and vaccine coverage, with estimated cases and deaths from AZ vaccine-associated TTS

Model version 2 was used to estimate COVID-19 deaths prevented over 6 months per million population if 70% had a first dose, and 35% had two doses. Figures 3a, 3b, and 3c show the estimated deaths prevented for each age group under different levels of community transmission:

**Figure 3.**
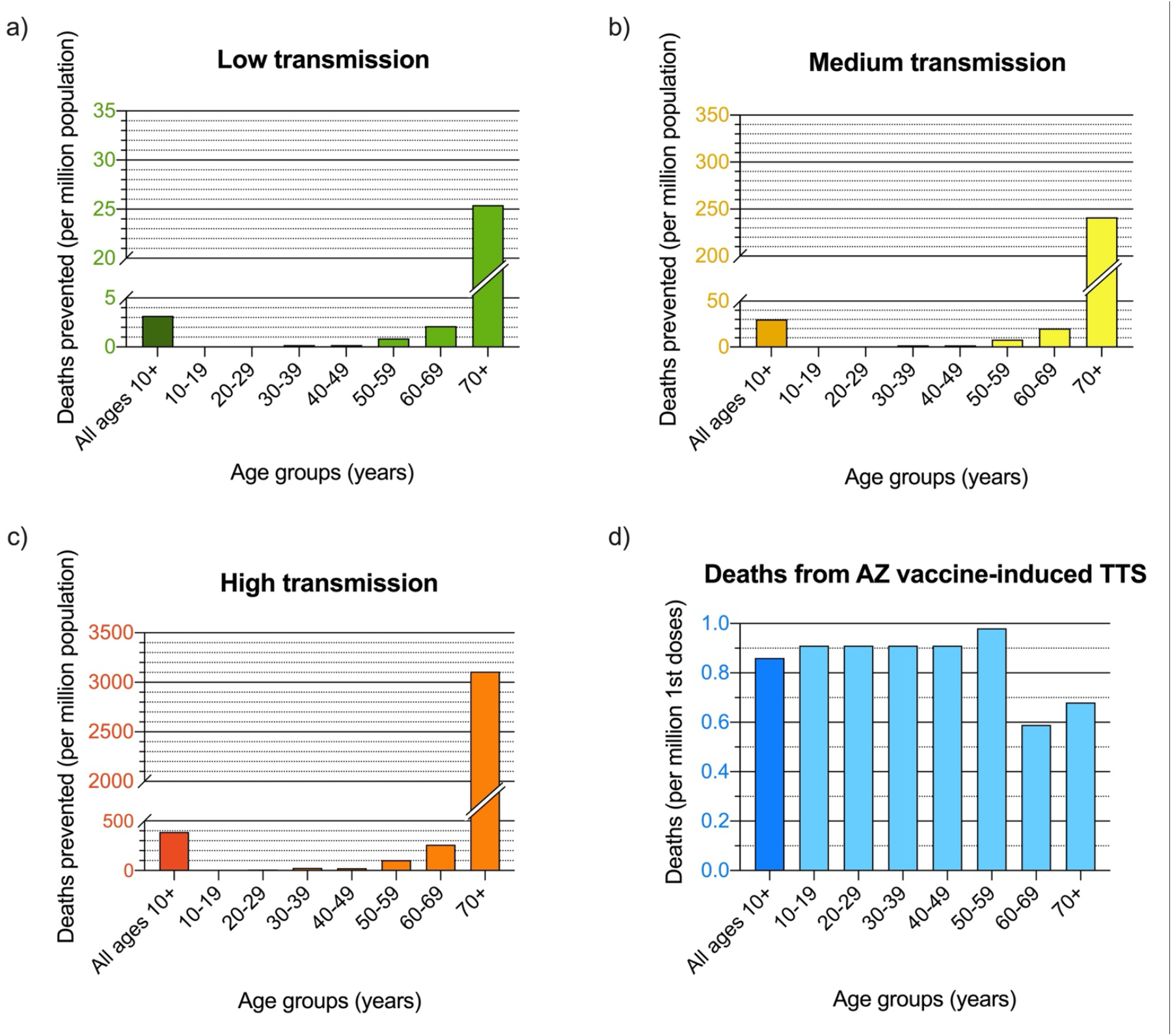
Estimated COVID-19 deaths prevented over 6 months per million population of each age group if 70% had first dose, and 35% had two doses of AZ vaccine under a) low, b) medium, and c) high levels of community transmission; and d) estimated deaths from AZ vaccine-associated TTS if 70% of the population had first dose, and 35% had two doses. (Note the large variations in scale in y-axes between each graph).

- Low transmission (Figure 3a), similar to 1^st^ wave in Australia in 2020, equivalent to 0.05% of population infected over 6 months;
- Medium transmission (Figure 3b), similar to 2^nd^ wave in VIC in 2020, equivalent to 0.45% of population infected over 6 months; and
- High transmission (Figure 3c), similar to Europe in January 2021, equivalent to 5.76% of population infected over 6 months.

The model shows that for a million people aged ≥70 years where 70% have had first dose and 35% two doses, an estimated 25 deaths would be prevented under low transmission (Fig 3a) versus >3000 deaths prevented under high transmission (Fig 3c), with <1 expected death from TTS (Fig 3d). In a million people aged 60-69 years with the same vaccine coverage, the model estimates two deaths prevented under low transmission, and 260 deaths prevented under high transmission, with <1 expected death from TTS. In contrast, for a million 20-29 year-olds with the same vaccine coverage, <0.1 deaths would be prevented under low transmission, ∼9 deaths prevented under high transmission, with <1 expected death from TTS. Details on calculations are provided in Appendix C.

### 3.4. Comparison of risk of AZ vaccine-associated TTS with risk of atypical blood clots (CVST and PVT) in COVID-19 infected patients

Up to 25/8/2021 in Australia, age-specific incidence of AZ vaccine-associated TTS cases ranged from 16 to 27 per million first doses (Table 1), with an overall CFR of ∼5% (6 deaths out of 115 cases). Based on annual background rates of CVST and PVT reported by Kristoffersen *et al*. [27] and Ageno *et al*. [28], our model estimated 6-week incidence of 0.38 (age <20 years) to 2.69 (age ≥70 years) cases per million, and overall CFR ranging from 7.0% to 21.6% from youngest to oldest age groups (Table A7). The background CFR for CVST and PVT (combined) range from 0.03 (age <20 years) to 0.58 (age ≥70 years) per million over 6 weeks, compared to CFR of 0.83 to 1.40 per million first doses of AZ vaccine depending on age (Table 3).

**Table 3.**
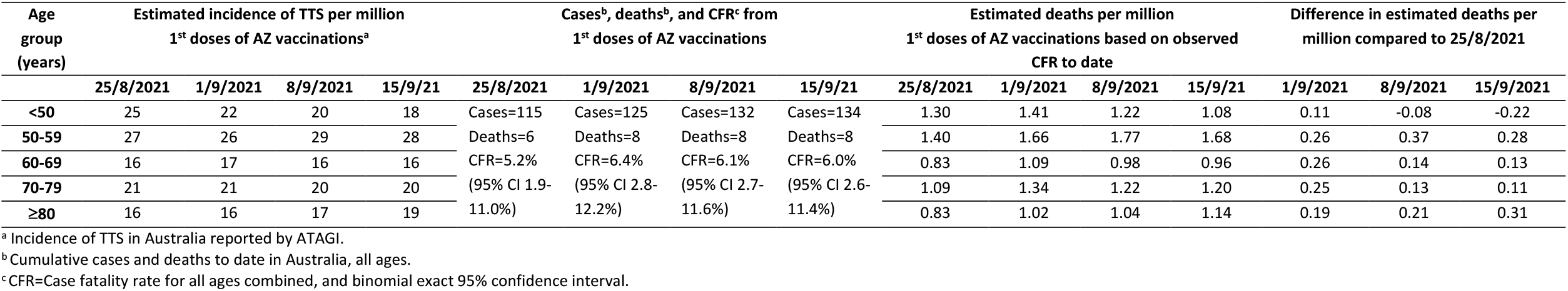
Evolving evidence on incidence and case-fatality rate (CFR) of vaccine-associated Thrombosis with Thrombocytopenia Syndrome (TTS) in Australia in August-September 2021, and influence on estimated TTS-related deaths by age group.

Model assumptions on the incidence and CFR of CVST and PVT in COVID-19 patients were obtained from a retrospective cohort study using linked electronic health records predominantly from the USA and Europe [30] because there were insufficient data from Australia. Model version 1 estimated that overall fatality from atypical severe blood clots (CVST and PVT combined) in COVID-19 patients were 51.1 and 37.3 per 100,000 in males and females, respectively. Figure 4 shows that the probability of developing atypical blood clots in COVID-19 patients was 14-28 times more likely than developing TTS after the first dose of the AZ vaccine, depending on age group and sex (Figure 4, dashed lines). The probability of dying from COVID-19 infection-related atypical blood clots in COVID-19 patients was 58-126 times more likely than dying from TTS after the first dose of AZ vaccine, again depending on age group and sex (Figure 4, solid lines).

**Figure 4.**
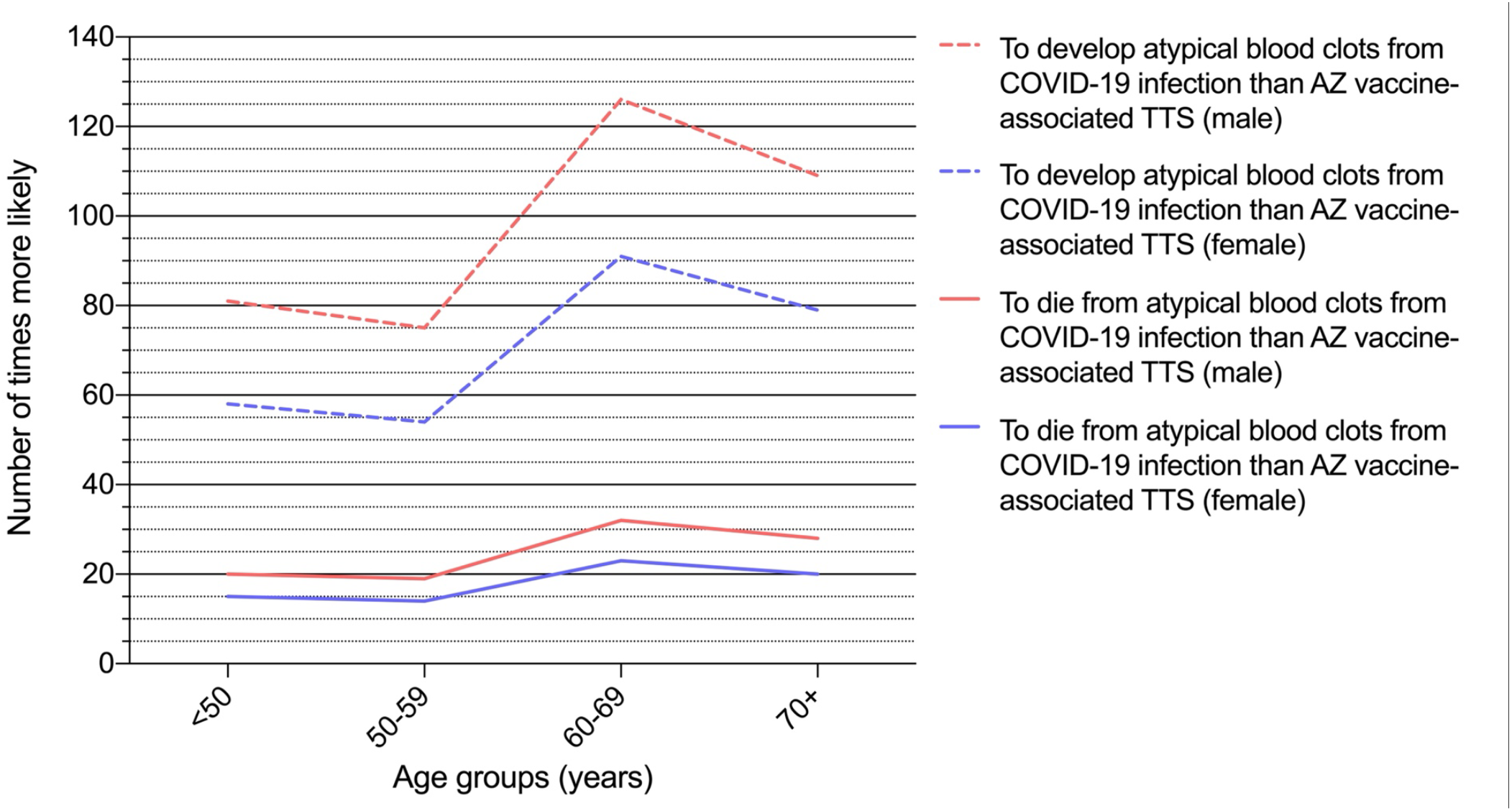
Number of times more likely to develop and die from atypical blood clots (CVST and PVT) from COVID-19 infection than AZ vaccine-associated TTS, by age group and sex.

### 3.5. Sensitivity analysis

#### Incidence and CFR of AZ vaccine-associated TTS

ATAGI reports from 25/8/2021 to 15/9/2021 [36-39] showed minor changes in the incidence of vaccine-associated TTS in Australia, and CFR ranging from 5.2% to 6.4% (Table 3). The model outputs showed that estimated deaths from TTS per million first doses did not change significantly over this time period, and ranged from differences of -0.08 to 0.37 deaths per million (depending on age group) when comparing data from 25/8/2021 with subsequent reports. Therefore, minor fluctuations in CFR from TTS did not have any significant influence on the point estimates of the number deaths at a population level. Because of the small number of TTS cases and deaths in Australia so far, the 95% confidence intervals (CI) for CFR were wide, ranging from 1.9% to 12.2% over the four reports (Table 3). Our model currently assumes a CFR of 5%; it is plausible that true CFR is twice as high (10%), and if so, model estimates of TTS related deaths would be doubled.

#### Age-specific CFR for COVID-19 in Australia

By August 2021, CFR for COVID-19 in Australia was 1.84% in males and 1.94% in females. In those aged under 50 years, CFR was <0.1% in both sexes (Appendix A). If there was one extra death in each age-sex subgroup during this time, CFR would have increased the most in 60-69 year-olds and the least in 20-29 year-olds, but by less than 0.06% in any subgroup (Figure 5). The model was most sensitive to changes in the 60-69 year-old age group because of the small case numbers (small denominator). If there were five extra deaths in each age-sex subgroup, CFR would have increased by less than 0.3% in any subgroup. Therefore, our model was not very sensitive to minor changes in number of reported deaths, and model estimates of deaths per million would have differed by less than 0.06% or 0.3% if there were one or five extra deaths in any age-sex subgroup, respectively.

**Figure 5.**
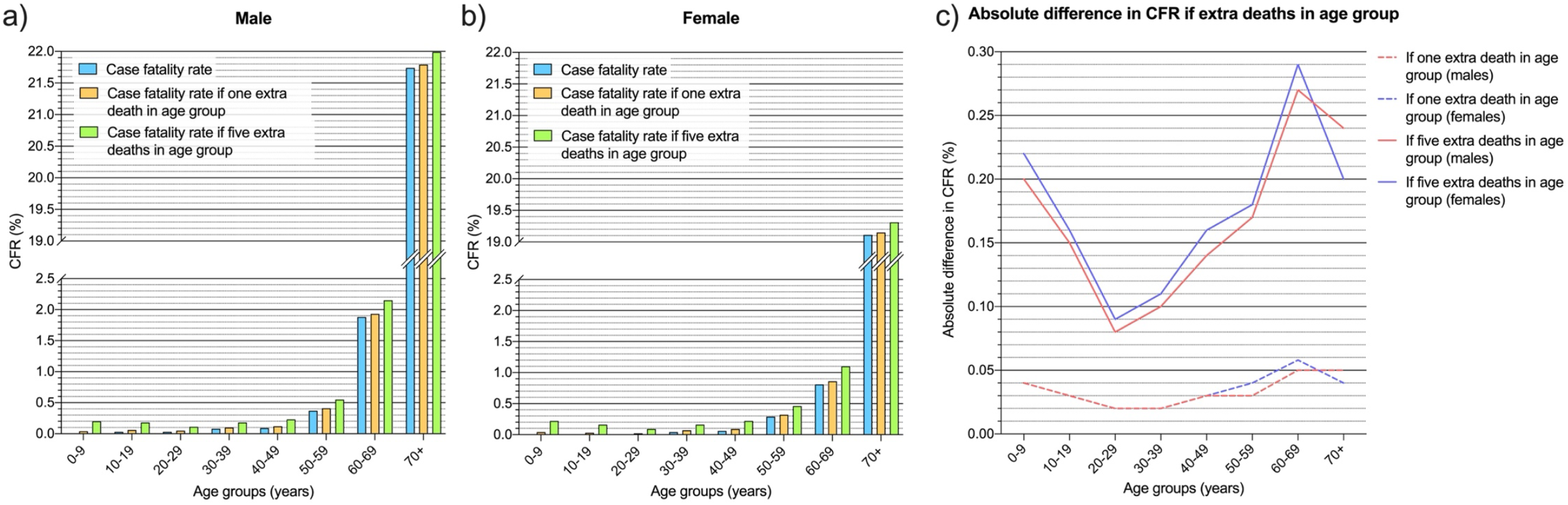
Change in age-sex specific case fatality rate for COVID-19 if one or five extra deaths in each subgroup: a) CFR in males, b) CFR in females, c) absolute difference in CFR if one or five extra deaths. (Based on COVID-19 cases and deaths in Australia by 31/8/2021).

#### Vaccine effectiveness against symptomatic infection and death

Our model shows that for a population where 70% has received first dose and 35% has received two doses, a theoretical 5% or 10% reduction in vaccine effectiveness against the delta variant results in a 7.1% or 15.1% increase in estimated deaths, respectively (Table 4). Therefore, sensitivity analyses show that model predictions of deaths are much more sensitive to changes in vaccine effectiveness than to changes in incidence and CFR of TTS, or changes in CFR from COVID-19 infection.

**Table 4.** Impact of theoretical reduction in vaccine effectiveness against delta variant on estimated deaths, assuming 70% of population had first dose, and 35% had two doses.

| Delta variant | Current model assumptions | If 5% less effective | If 10% less effective |
| --- | --- | --- | --- |
| Vaccine effectiveness against symptomatic infection after |  |  |  |
| • 1<sup>st</sup> dose | 33% | 28% | 23% |
| • 2<sup>nd</sup> dose | 61% | 56% | 51% |
| Vaccine effectiveness against death after |  |  |  |
| • 1<sup>st</sup> dose | 69% | 64% | 59% |
| • 2<sup>nd</sup> dose | 90% | 85% | 80% |
| % Increase in estimated deaths compared to current model assumptions of vaccine effectiveness | N/A | 7.1% | 15.1% |

## 4. DISCUSSION

Our model demonstrates that risk-benefit analysis of the AZ vaccine is complex, and depends on multiple factors including age, sex, vaccine effectiveness, and locally specific factors such as the intensity of community transmission, local incidence and CFR of TTS, and CFR from COVID-19. The model outputs and scenario analysis could facilitate more informed decision making between clinicians and individuals, e.g., by considering age, sex, community transmission, and local data on TTS. The model could also provide decision support at a public health level; if the priors were set to represent the population in age and sex distribution, vaccination coverage, circulating variants and level of community transmission, the model outputs could be used to estimate deaths or deaths prevented. To our knowledge, this is the first example of the use of Bayesian networks for risk-benefit analysis for a COVID-19 vaccine.

Our model quantifies the risks (deaths from TTS) versus benefits from vaccination (deaths prevented) for different age groups under different levels of community transmission (Figure 3). While the benefits are significantly greater during higher transmission (especially for older age groups), decisions about vaccination should be based not only on the current level of transmission, but also potential future scenarios. For example, higher transmission will be almost inevitable with lifting of public health restrictions and opening of borders. We presented results for scenarios where 70% of the population have had first dose and 35% have had two doses. Other vaccination coverage scenarios can be easily defined in the model priors to assess the current vaccination coverage rates, or future vaccination targets.

To date, the CFR of TTS in Australia has been lower than those reported in other countries [7, 9]. Although numbers have been small, CFR have remained relatively stable throughout August-September 2021 (Table 3). Possible reasons for the lower CFR in Australia include early shift to Pfizer as the preferred vaccine for those aged <60 years; detailed expert review of each case of serious and fatal AEFI (reducing classification error) by state-based committees, ATAGI and TGA; high clinical vigilance of TTS based on lessons learnt from earlier reports from other countries; and clinical guidelines for vaccine providers on early diagnosis and referral of possible TTS cases [41-43]. Our sensitivity analysis showed that fluctuations in observed CFR from TTS had only minor impact on model outputs. Model assumptions can be readily updated if CFR changes significantly in the future. While vaccine-associated TTS of any type can be serious and rarely fatal, mortality for those presenting with atypical severe blood clots (CVST and PVT) in COVID-19 patients are estimated to be 58-126 times more likely than after the first dose of the AZ vaccine (Figure 4).

Sensitivity analyses showed that the estimates of COVID-19 related deaths were very sensitive to changes in vaccine effectiveness. Therefore, model assumptions need to be updated to reflect evolving evidence on vaccine effectiveness against current and future variants of the virus, waning immunity over time, and effectiveness of vaccine boosters. From a public health perspective, our findings suggest that a decrease in vaccine effectiveness will likely have important implications on disease burden. In contrast, model outputs were not very sensitive to changes in age-specific CFR for COVID-19, and updates to model assumptions are unlikely to be required unless changes are dramatic.

We used an innovative and flexible modelling framework which allows model assumptions to be easily changed, either to reflect new data (e.g., vaccine effectiveness), to the evolving situation (e.g., intensity of community transmission, changing CFR from TTS and COVID-19). While the current model is designed to estimate deaths from vaccine versus disease, it could be readily adapted for other outcomes (e.g., ICU admission, long COVID), other adverse events (e.g., immune thrombocytopenia), risk-benefit assessment of other vaccines (e.g., myocarditis/pericarditis from mRNA vaccines), and other types of scenario analysis (e.g., different combinations of vaccine doses and boosters). Our modelling approach also enables the use of multiple sources of data. We have used Australian data wherever possible and international sources if needed (e.g., low number of COVID-19 cases and vaccinations in Australia means that international data on vaccine effectiveness were likely to be more accurate). Our modelling approach therefore provides the potential for countries to develop a locally relevant risk-benefit assessment tool for COVID-19 vaccination even if there are limited local data.

Another advantage of BNs is the visual and interactive interface that allows intuitive scenario analysis for users and decision makers. This paper presents results of specific scenarios from our model, but we aim to produce interactive versions of the model and make them freely available on the Immunisation Coalition website (https://www.immunisationcoalition.org.au/). Interactive risk assessment tools for COVID-19 have been developed for estimating the risk of hospitalization and death (e.g., QCOVID tool in United Kingdom [44], COVID-19 risk tools in USA [45] and France [46], predicting clinical outcomes [47], and assessing risk of infection from different activities [48-50]. Some tools were based on analysis of millions (e.g., QCOVID [44]) or hundreds of thousands (e.g., ISARIC4C [47]) of medical records. Our modelling approach provides an alternative option of developing risk assessment tools if large datasets are not available, and model inputs need to be sourced from alternative sources. We have not identified any web-based interactive tools that use Bayesian networks for risk-benefit analysis of COVID-19 vaccines for use in clinical or public health settings.

Our results should be interpreted considering the model’s limitations. There are uncertainties associated with some of our model inputs, either because of limited data, or the use of data from other countries. As illustrated with the anomaly regarding the risk of dying in younger persons, model outputs should be considered as broad estimates rather than exact risks, but estimates can be improved over time as more data become available. Our model provides population level estimates and does not consider individual risks such as behaviour and comorbidities. We plan to develop future models that include the individual’s comorbidities, similar to the QCOVID tool [44] but specific for the Australian context. In the results provided, we have assumed that 100% of infections were from the delta variant. Assumptions of age distribution of delta cases (if unvaccinated) were obtained from data during the early stages of the delta outbreak in NSW from June 2021. While vaccination rates were relatively low then, older ages had higher vaccine coverage so infection rates for delta may have been underestimated in these groups. Data on CVST and PVT were obtained from studies outside Australia and may not reflect the local experience. The current model focuses on fatalities from COVID-19, TTS, and atypical blood clots, but does not consider other risks (e.g., adverse events) or other benefits (e.g., cases of severe COVID prevented, or broader societal benefits). Our model was not parameterised from any specific datasets, so model outputs could not be directly validated by data. Nevertheless, the model provides a powerful mechanism for complex synthesis of multiple sources of data, and the outputs reflect the latest available knowledge.

In conclusion, we developed a novel approach to risk-benefit analysis for the AZ vaccine by using an adaptable BN modelling framework. Our model enables more precise risk analysis based on demographics, the outbreak situation, local data on vaccine-associated TTS, and the best available international evidence on vaccine effectiveness and atypical blood clots. Although use of the AZ vaccine is expected to gradually decrease in Australia over coming months, the model can be readily adapted for use in other countries or risk-benefit assessment of other vaccines.

## Data Availability

All data referred to in the manuscript are accessible online.

## Author contributions

Conception and design: JL, AB, KRS, CLL, HJM, KM

Acquisition of data: JL, AB, KRS, CLL, HJM, JES, SJB, AKE

Analysis and interpretation: CLL, HJM, KM, JL, JES, SJB, MW

Drafting the article: CLL

Revising article for important intellectual content: All authors

Final approval of submitted version: All authors

## Acknowledgements

We thank Kim Sampson from Immunisation Coalition for facilitating the collaboration between authors, and A/Prof Hassan Valley (La Trobe University, Melbourne, Australia) for contributions to discussions about risk communication and data visualisation. We thank Aapeli Vuorinen (Columbia University) for building the interactive risk assessment tool. Our Bayesian network model was built using GeNIe Modeler (BayesFusion 2019), available free of charge for academic research and teaching use from https://www.bayesfusion.com/. We thank the THANZ Vaccine-induced Immune Thrombotic Thrombocytopenia (VITT) advisory group (in particular A/Prof Vivien Chen, Prof Huyen Tran, A/Prof Jennifer Curnow, Dr Ibrahim Tohidi and Dr Danny Hsu) for their feedback about TTS related data.

