## Appendix for "Risk-benefit analysis of the AstraZeneca COVID-19 vaccine in Australia using a Bayesian network modelling framework"

### Appendix A. Model assumptions

**Table A1. Age distribution of reported cases of COVID-19 in NSW, 29/6/2021 to 13/8/2021.**

| Age group | Cases |
|---|---|
| 0-9* | 734 |
| 10-19* | 1100 |
| 20-29 | 1399 |
| 30-39 | 1064 |
| 40-49 | 752 |
| 50-59 | 710 |
| 60-69 | 371 |
| ≥70 | 319 |
| **Total** | **6449** |

*NSW Health reported 1834 cases in the 0-19 age group. Numbers based on assumption that 40% cases in the 0-19 age group were aged 0-9, and 60% were aged 10-19 age.

Sources:

NSW Government *NSW COVID-19 cases data*. 2021. https://data.nsw.gov.au/nsw-covid-19-data/cases. (31)

Australian Government Department of Health *Cases and deaths by age and sex*. Coronavirus (COVID-19) case numbers and statistics, 2021. https://www.health.gov.au/news/health-alerts/novel-coronavirus-2019-ncov-health-alert/coronavirus-covid-19-case-numbers-and-statistics#cases-and-deaths-by-age-and-sex. (32)

**Table A2. Age distribution of reported cases of COVID-19 in Australia, January to December 2020.**

| Age group | Cases |
|---|---|
| 0-9 | 1480 |
| 10-19 | 2416 |
| 20-29 | 6419 |
| 30-39 | 5072 |
| 40-49 | 3687 |
| 50-59 | 3417 |
| 60-69 | 2445 |
| ≥70 | 3713 |
| **Total** | **28649** |

Sources:

Australian Government Department of Health *COVID-19 summary statistics*. Coronavirus (COVID-19) case numbers and statistics, 2021. https://www.health.gov.au/news/health-alerts/novel-coronavirus-2019-ncov-health-alert/coronavirus-covid-19-case-numbers-and-statistics#covid19-summary-statistics. (14)

Australian Government Department of Health *National Notifiable Diseases Surveillance System public datasets*. 2021. https://www1.health.gov.au/internet/main/publishing.nsf/Content/ohp-pub-datasets.htm. (33)

**Table A3. Cases, deaths, and case fatality rate of COVID-19 in Australia by age and gender, January 2020 to 31/8/2021.**

| | Male | | | Female | | |
|---|---|---|---|---|---|---|
| **Age group** | **Cases** | **Deaths** | **Case fatality** | **Cases** | **Deaths** | **Case fatality** |
| 0-9 | 2848 | 0 | 0% | 2317 | 0 | 0% |
| 10-19 | 3298 | 1 | 0.03% | 3196 | 0 | 0% |
| 20-29 | 6162 | 2 | 0.03% | 5859 | 0 | 0% |
| 30-39 | 4975 | 4 | 0.08% | 4511 | 2 | 0.04% |
| 40-49 | 3468 | 3 | 0.09% | 3178 | 2 | 0.06% |
| 50-59 | 2935 | 11 | 0.37% | 2798 | 8 | 0.29% |
| 60-69 | 1862 | 35 | 1.88% | 1735 | 14 | 0.81% |
| ≥70 | 2042 | 444 | 21.74% | 2512 | 480 | 19.11% |
| **Total** | **27,226** | **500** | **1.84%** | **26,106** | **506** | **1.94%** |

Sources:

Australian Government Department of Health *COVID-19 summary statistics*. Coronavirus (COVID-19) case numbers and statistics, 2021. https://www.health.gov.au/news/health-alerts/novel-coronavirus-2019-ncov-health-alert/coronavirus-covid-19-case-numbers-and-statistics#covid19-summary-statistics. (14)

Australian Government Department of Health *National Notifiable Diseases Surveillance System public datasets*. 2021. https://www1.health.gov.au/internet/main/publishing.nsf/Content/ohp-pub-datasets.htm. (33)

**Table A4. Probability of infection (over 6 months) based on different intensities of community transmission.**

| **Intensity of community transmission** | **Cases per 100,000 over 16 weeks*** | **Cases per million over 6 months** | **Estimated % of population infected over 6 months** |
|---|---|---|---|
| Zero | 0 | 0 | 0% |
| Low* | 29 | 471 | 0.047% |
| Medium* | 275 | 4,469 | 0.447% |
| High* | 3,544 | 57,590 | 5.759% |
| 1% over 6 months | | 10,000 | 1.0% |
| 2% over 6 months | | 20,000 | 2.0% |
| 200 cases/day in NSW[a] | | 4,455 | 0.446% |
| 1000 cases/day in NSW[a] | | 22,276 | 2.228% |
| 1000 cases/day in VIC[b] | | 27,289 | 2.729% |
| 1000 cases/day in QLD[c] | | 35,067 | 3.507% |

* Definitions of low, medium, and high transmission (cases per 100,000 over 16 weeks) as defined by ATAGI document 'Weighing up the potential benefits and risk of harm from COVID-19 Vaccine AstraZeneca'. Low – similar to first wave in Australia. Medium – similar to second wave in VIC. High – similar to Europe in January 2021.

[a]Based on NSW population of 8.17 million. [b]Based on VIC population of 6.67 million. [c]Based on QLD population of 5.19 million.

Source:

Australian Government Department of Health *COVID-19 vaccination – Weighing up the potential benefits against risk of harm from COVID-19 Vaccine AstraZenec*. 2021. https://www.health.gov.au/resources/publications/covid-19-vaccination-weighing-up-the-potential-benefits-against-risk-of-harm-from-covid-19-vaccine-astrazeneca. (15)

**Table A5. Relative risk of infection by age group and variant (chance of infection if overall probability of infection of 1% in all ages).**

| Age group | Alpha/wild | Delta |
|---|---|---|
| 0-9 | 0.42% | 0.92% |
| 10-19 | 0.70% | 1.42% |
| 20-29 | 1.59% | 1.54% |
| 30-39 | 1.21% | 1.13% |
| 40-49 | 1.01% | 0.91% |
| 50-59 | 0.98% | 0.90% |
| 60-69 | 0.81% | 0.55% |
| ≥70 | 1.13% | 0.43% |
| **Overall** | **1.00%** | **1.00%** |

**Table A6. Age distribution of Australian population, December 2020.**

| Age group | Population | % of total population |
|---|---|---|
| 0-9 | 3,189,750 | 12.4% |
| 10-19 | 3,086,855 | 12.0% |
| 20-29 | 3,627,055 | 14.1% |
| 30-39 | 3,755,673 | 14.6% |
| 40-49 | 3,292,645 | 12.8% |
| 50-59 | 3,138,303 | 12.2% |
| 60-69 | 2,700,998 | 10.5% |
| ≥70 | 2,958,236 | 11.5% |
| **Total** | **25,749,515** | **100%** |

Source:
Australian Bureau of Statistics *Data downloads - time series spreadsheets*. National, state and territory population, 2021. https://www.abs.gov.au/statistics/people/population/national-state-and-territory-population/dec-2020#data-download. (40)

**Table A7. Estimated background incidence and fatality of atypical blood clots (CVST and PVT combined) over 6 weeks (in populations who have not received AZ vaccine and not diagnosed with COVID-19).**

| Age group | Incidence of atypical blood clots (CVST and PVT) over 6 weeks (per million) | Fatality rates from atypical blood clots (CVST and PVT) over 6 weeks (per million) | Overall case fatality rate from atypical blood clots (CVST and PVT) |
|---|---|---|---|
| 0-9 | 0.38 | 0.03 | 7.0% |
| 10-19 | 0.38 | 0.03 | 7.0% |
| 20-29 | 0.84 | 0.10 | 11.8% |
| 30-39 | 0.90 | 0.12 | 12.8% |
| 40-49 | 1.20 | 0.20 | 16.3% |
| 50-59 | 1.66 | 0.30 | 18.0% |
| 60-69 | 2.51 | 0.53 | 21.0% |
| ≥70 | 2.69 | 0.58 | 21.6% |

**Appendix B.**

**Examples of scenarios used for manual model validation.**

Assume transmission of delta variant only for all scenarios.

1. For a 30-39 year-old male, what is the chance of symptomatic infection under high transmission if: a) Not vaccinated, b) had one dose of AZ vaccine, c) had two doses of AZ vaccine.
2. or a 30-39 year-old male, what is the overall chance of dying under high transmission if: a) Not vaccinated, b) had one dose of AZ vaccine, c) had two doses of AZ vaccine.
3. For a million 50-59 year-old females, if 70% had first dose, and 35% had two doses: a) How many cases of vaccine induced TTS would we expect? b) How many TTS related deaths would we expect?
4. For a million 70+ year-old males, if 70% had first does, and 35% had two doses: a) How many symptomatic cases would we expect over 6 months if there was ATAGI medium transmission during this time? B) How many deaths would we expect over 6 months if there was ATAGI medium transmission during this time?
5. If a 60-69 year-old female was diagnosed with COVID-19: a) what are her chances of developing atypical blood clots (CVST and PVT)? b) what are her chances of dying from atypical blood clots (CVST and PVT)?

**Appendix C.**

**Table C1. Comparison of deaths prevented by AstraZeneca COVID-19 vaccine vs deaths from vaccine-associated Thrombosis with Thrombocytopenia Syndrome (TTS) under different intensities of community transmission.** Assuming transmission of delta variant; 30% of unvaccinated, 70% received first dose, 35% received two doses; vaccine effectiveness against death of 69% after 1st dose, 90% after 2nd dose; age-specific TTS incidence and case-fatality rate (5%) reported by ATAGI on 25th August 2021.

| Age group (years) | Community transmission intensity[a] (probability of infection over 6 months) | Estimated COVID-19 deaths over 6 months (per million[b]) | | Estimated COVID-19 deaths prevented over 6 months if 70% had 1st dose, 35% had 2 doses (per million[b]) | Estimated cases of vaccine-associated TTS if 70% had 1st dose, 35% had 2 doses (per million[b]) | Estimated deaths from vaccine-associated TTS if 70% had 1st dose, 35% had 2 doses (per million[b]) | Estimated deaths prevented per vaccine-associated death from TTS |
|---|---|---|---|---|---|---|---|
| | | 0% vaccinated | 70% had 1st dose, 35% had 2 doses | | | | |
| All ages ≥10[c] | Low (0.047%) | 5.2 | 2.0 | 3.2 | 17.2 | 0.9 | 3.7 |
| | Medium (0.447%) | 49.2 | 19.0 | 30.2 | | | 35.1 |
| | High (5.76%) | 634.3 | 245.1 | 389.3 | | | 452.6 |
| 10-19 | Low (0.047%) | <0.1 | <0.1 | <0.1 | 18.1 | 0.9 | 0.1 |
| | Medium (0.447%) | 1.0 | 0.4 | 0.6 | | | 0.7 |
| | High (5.76%) | 12.4 | 4.8 | 7.6 | | | 8.4 |
| 20-29 | Low (0.047%) | 0.1 | <0.1 | <0.1 | 18.1 | 0.9 | 0.1 |
| | Medium (0.447%) | 1.1 | 0.4 | 0.7 | | | 0.8 |
| | High (5.76%) | 14.4 | 5.6 | 8.8 | | | 9.7 |
| 30-39 | Low (0.047%) | 0.3 | 0.1 | 0.2 | 18.1 | 0.9 | 0.2 |
| | Medium (0.447%) | 3.2 | 1.2 | 1.9 | | | 2.1 |
| | High (5.76%) | 40.6 | 15.7 | 24.9 | | | 27.4 |
| 40-49 | Low (0.047%) | 0.3 | 0.1 | 0.2 | 18.1 | 0.9 | 0.2 |
| | Medium (0.447%) | 3.1 | 1.2 | 1.9 | | | 2.1 |
| | High (5.76%) | 39.2 | 15.2 | 24.1 | | | 26.5 |
| 50-59 | Low (0.047%) | 1.4 | 0.5 | 0.9 | 19.5 | 1.0 | 0.9 |
| | Medium (0.447%) | 13.3 | 5.2 | 8.2 | | | 8.4 |
| | High (5.76%) | 171.9 | 66.4 | 105.5 | | | 107.6 |
| 60-69 | Low (0.047%) | 3.5 | 1.3 | 2.1 | 11.8 | 0.6 | 3.6 |
| | Medium (0.447%) | 32.9 | 12.7 | 20.2 | | | 34.2 |
| | High (5.76%) | 424.3 | 163.9 | 260.4 | | | 441.3 |
| ≥70 | Low (0.047%) | 41.4 | 16.0 | 25.4 | 13.6 | 0.7 | 37.4 |
| | Medium (0.447%) | 393.0 | 151.8 | 241.2 | | | 354.7 |
| | High (5.76%) | 5064.8 | 1956.8 | 3108.0 | | | 4570.6 |

[a] Definitions of community transmission intensity based on ATAGI document 'Weighing up the potential benefits and risk of harm from AstraZeneca COVID-19 vaccine. Low transmission: similar to first wave in Australia in 2020 (equivalent to 0.05% of population infected over 6 months). Medium transmission: similar to second wave in Victoria, Australia in 2020 (equivalent to 0.45% of population infected over 6 months). High transmission: similar to Europe in January 2021 (equivalent to 5.76% of population infected over 6 months).

[b] Per million population of each age group, or per million of all ages based on population distribution of Australia

[c] Calculations for all ages based on population distribution of Australia
